# The effect of including the Nordic Hamstring exercise on sprint and jump performance in athletes: protocol of a systematic review and meta-analyses

**DOI:** 10.1101/2020.04.01.20048686

**Authors:** Kasper Krommes, Mathias F. Nielsen, Laura Krohn, Birk M. Grønfeldt, Kristian Thorborg, Per Hölmich, Lasse Ishøi

## Abstract

The Nordic Hamstring exercise reduces hamstring strain injuries in football and other sports, but the exercise is not well adopted in practice. Barriers from practitioners include fear of performance decrements, due to lack of specificity of the exercise with high speed running. However, in theory, increased eccentric hamstring strength could transfer to faster sprinting due to higher horizontal force production. Studies on the effect of the Nordic Hamstring exercise on performance have been conflicting and no synthesis of the evidence exists. We therefore pose the following question: does including the Nordic Hamstring exercise hamper sprint or jump performance in athletes? We will answer this question by performing a systematic review of the literature, critically appraise relevant studies, and GRADE the evidence across key outcomes and perform meta-analyses, meta-regression and subgroup analyses. In this protocol we outline the planned methods and procedures.

**Progress report:** Besides this protocol, our data extraction form and the process of data extraction has been piloted on 3 relevant studies, along with familiarization with the Risk of Bias 2.0 tool. We have also comprised a preliminary search strategy for PubMed.

**Supplementary files:**

- Data Extraction Form (.pdf)
- Populated PRISMA-P checklist (.pdf)

## 1.0 BACKGROUND

### 1.1 Introduction

Exercise and sports participation benefits health,^1–3^ all-cause mortality^4–6^ and disease prevention.^7–10^ Football and its various codes are the most popular sports worldwide,^11^ but do carry risk of injuries,^12^ of which the most common and burdensome are hamstring strain injuries.^13–15^ These injuries are preventable by implementation of the Nordic Hamstring exercise in various doses,^16^ even in other sports besides football.^17–19^ However, even at the elite football level the exercise is not implemented,^20^ and hamstring strain injuries continue to top the injury-statistics.^21^ Narratives and surveys from the Sports Medicine community suggests that fear of performance-decrements and soreness from doing the exercise are barriers to implementation.^22–25^ Recently, randomized controlled trials from elite and amateur football and handball players have shown improvements in sprint performance after Nordic hamstring exercise-interventions,^26–28^ although the mechanism responsible has not yet been established.^29^ The most relevant performance measure in football and other sports is sprint ability,^30,31^ however, potential sprint performance effects of doing the Nordic Hamstring exercise has not yet been synthesized across studies. Such a synthesis address some of the concerns and barriers for implementation of the Nordic Hamstring exercise and supply new data-based arguments regarding impacts on performance, and in consequence likely increase adoption.

### 1.2 Objective and research questions

In order to provide a data-based estimate on the effect of including the Nordic Hamstring exercise in the team warm-up or in conditioning regiment on performance measures, we aim to answer the following questions:

#### Primary question

What is the effect of including the Nordic Hamstring exercise on sprint performance in athletes?

#### Secondary question

What is the effect of including the Nordic Hamstring exercise on other performance measures (such as: change of direction/agility, jumping, repeated sprint performance); and does it carry a risk of adverse events (soreness, injuries, and any other reported adverse events)?

## 2.0 METHODS

The protocol for this review will adhere to the PRISMA-P (Preferred Reporting Items for Systematic reviews and Meta-analyses - Protocol) statement^32^ and the proposed search-extension (PRISMA-S)^33^ to increase transparency and reproducibility,^34^ and we aim to fulfil items on AMSTAR-2 (A MeaSurement Tool to Assess systematic Reviews)^35^ to increase study quality, and ROBIS (Risk Of Bias In Systematic reviews)^36^ to minimize risk of bias. Additionally, the final report will adhere to the PRISMA statement and the extension for abstracts.^37,38^Along with this pre-printed protocol, we have registered the study in the PROSPERO (international PROSPEctive Register Of systematic reviews) repository before commencement of data collection to adhere to *a priori* decisions (submitted 31-MAR-2020, identifier pending).^36,39^ After a duration of public peer-review as pre-print, a possible updated version of the protocol will be uploaded before we commence data collection and perform literature searches, along with an update to the PROSPERO registration. After publication, all data (populated data extraction forms, search files, supplementary analyses, statistical code, bias- and GRADE (Grading of Recommendations Assessment, Development and Evaluation) assessments, populated PRISMA checklist) will be shared as supplementary files with no restrictions, either on our institutional website, per journal repository or other open access repository (e.g. figshare.com).

### 2.1 Outcomes

Modern football and other field-based sports is characterized by large amounts of sprint acceleration efforts,^30^ and consequently exercise interventions to improve this are considered a high priority.^40,41^ A decrease of 0.05 s in a 10 meter sprint time by an average player/athlete, may likely result in a 25–30 cm lead during a maximal 10 m sprint.^42^ In e.g. football, such a difference is considered clinically important, and may be crucial to reach a ball before the opponent player, block a shot, or score a goal.^42^ Most sprints in football are under 20 meters, ^43^ and while sprinting, team-sport athletes plateau in velocity after approximately 20 meters^44^ and thus acceleration will be classified as sprinting distances <20 meters and maximal velocity as sprinting distances ≥20 meters. To obtain low measurement error, we will only consider sprint times measured by sensor-based systems, such as high-speed video, laser-sensors, radar-gun, or force-plates.

#### Primary outcome

- Change in sprint performance (seconds) during the phase of primarily acceleration (<20 meters)

#### Secondary outcomes (in prioritized order)

- Change in sprint performance (seconds) during combined acceleration/maximal velocity phase (≥20 meters)
- Changes in maximal velocity, measured as either velocity (m/s or km/h) or split times during maximal velocity running (between 20 to 40 meter)
- Change in repeated sprint ability
- Change in agility (such as: ‘change of direction’ test)
- Changes in jump performance (such as countermovement-jump, drop-jump, squat-jump etc.)
- Soreness or other adverse effects related to performing the Nordic Hamstring exercise

### 2.2 Criteria for eligibility of studies

#### 2.2.1 Outcomes of eligible studies

- Change in sprint time during acceleration phase (split times on 0 – <20 meters)
- Change in sprint time during acceleration and maximal velocity phase (split times on 0 - ≥20 meters)
- Change in maximal velocity (velocity or split times on ≥20 – 40 meters)
- Change in repeated sprint ability
- Change in agility
- Change in jump performance
- Soreness or other adverse effects related to performing the Nordic Hamstring Exercise

#### 2.2.2 Interventions of eligible studies

Studies prescribing the Nordic Hamstring exercise in any dose exceeding a minimum of 1 set of 3 repetitions (minimum session-dose in the fifa11+ injury prevention programmes^45^) per week in volume for a minimum of 3 weeks will be considered. The Nordic hamstring exercise should be performed either as originally described by Mjølsnes *et al*^46^ or with arms positioned in front of- or behind the body, or on devices substituting the partner, such as the ‘Nordbord’, ‘Hamstring Solo’ or other equipment. Additional loading during the exercise to maintain the supramaximal loading of the exercise is allowed, but not assisted variations that decrease the load to below bodyweight. Such variations will however be allowed if the programme also includes the conventional Nordic Hamstring exercise. The Nordic hamstring exercise can be performed in isolation as the only intervention, or as add-on to other types of exercise or conditioning. The Nordic Hamstring exercise and original protocol utilized in the literature is described in figure 1.^46^

**Figure 1.**
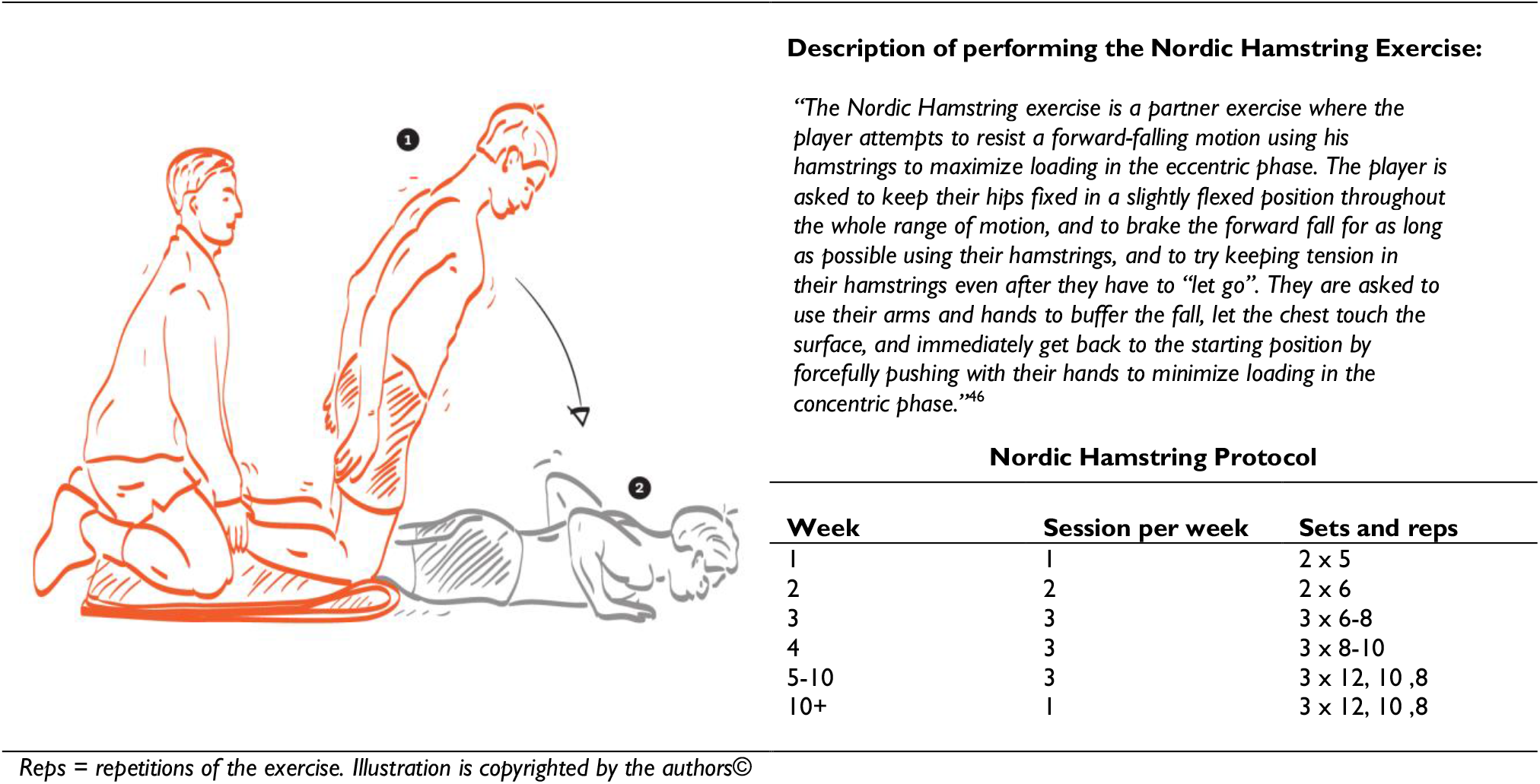
Description of the Nordic Hamstring Exercise and Protocol.

#### 2.2.3 Duration of follow-up in eligible studies

Studies doing follow-up testing 0 days to 4 weeks after termination of the intervention will be considered. For the acute effect of the Nordic Hamstring exercise, studies testing the same after doing the Nordic Hamstring Exercise will also be considered.

#### 2.2.4 Study design of eligible studies

All longitudinal studies implementing the Nordic Hamstring exercise will be considered, including single-group designs looking at within-group changes only, however, we will perform subgroup analyses for only RCTs and also for only low risk of bias RCTs.^47^ For the acute effect of the Nordic Hamstring exercise on sprint changes, cross-sectional studies with pre-post testing on the same day (effectively within-day longitudinal studies) will also be considered. Optimally, we would include only RCTs employing a non-inferiority framework, but given our knowledge of the field, such an evidence-pool is not available.

#### 2.2.5 Comparators of eligible studies

We will consider any comparators, such as usual training, no training, resistance training, specific sprint training or conditioning specifically for sprint performance enhancement. If studies have more than two groups we will use the Cochrane recommendations for deciding how to include data.^48^ For the primary analysis we will prioritize including usual care or similar comparators.

#### 2.2.6 Populations in eligible studies

Studies enrolling adult and adolescent participants (10-40 years) will be considered, whereas studies with children (<10 years) and older adults (>40 years) and the elderly will be excluded. Participants should at least be recreationally active, up to and including elite and professional athletes. We will perform subgroup analyses for adults or adolescents only (18-40 years, and 10-17 years), and for elite/professional athletes only.

### 2.3 Search strategy

We have consulted an expert football strength and conditioning coach at the elite level, and a research librarian for assistance in choosing terms and refining search strategy for the final search in PubMed, in addition to using the Word Frequency Analyser tool to suggest possible search terms from relevant literature (Systematic Review Accelerator, Institute for Evidence-Based Healthcare, Australia).^49^ We aim for a search strategy with high sensitivity and low precision. We have tested all terms individually, and some terms might be added or removed when applying the same strategy in other databases or platforms. Cochrane Central, Embase, PubMed and SPORTDiscus, will be sourced for relevant published studies,^50^ and the Polyglot Search Translator tool used to translated search strategies across databases (Systematic Review Accelerator, Institute for Evidence-Based Healthcare, Australia).^51^ In addition, any grey literature or unpublished data we can find through general search-engines (google.com, Google Scholar), forwards and backwards snowballing, trial-registries (ISRCTN, ICTRP, EU Register, OSF, ANZCTR), or by other means, will be sourced by hand search.^52^ The preliminary search matrix for PubMed is presented below (table 3), and will be adapted to all utilized databases to fit their respective search hierarchy, thesauruses, and operators. To our current knowledge of the field, titles and abstract does not always specify population or design fully thus we will not utilize search terms for these items, in order to not omit relevant studies. No filters or restrictions on dates, study design, population or language will be applied.^53^ If relevant articles with English abstracts but full-text language in other than Danish, Norwegian, Swedish or English languages are available, translation services will be sought. A copy of the search-results will be provided as supplementary files. The search process is planned for June 8^th^ – June 16^th^ 2020. For incomplete literature such as conference abstracts or theses, or insufficient reporting, we will contact corresponding authors to supply either the full report or information regarding risk of bias and GRADE assessment items, and to verify values for data extraction.

**Table 1.**
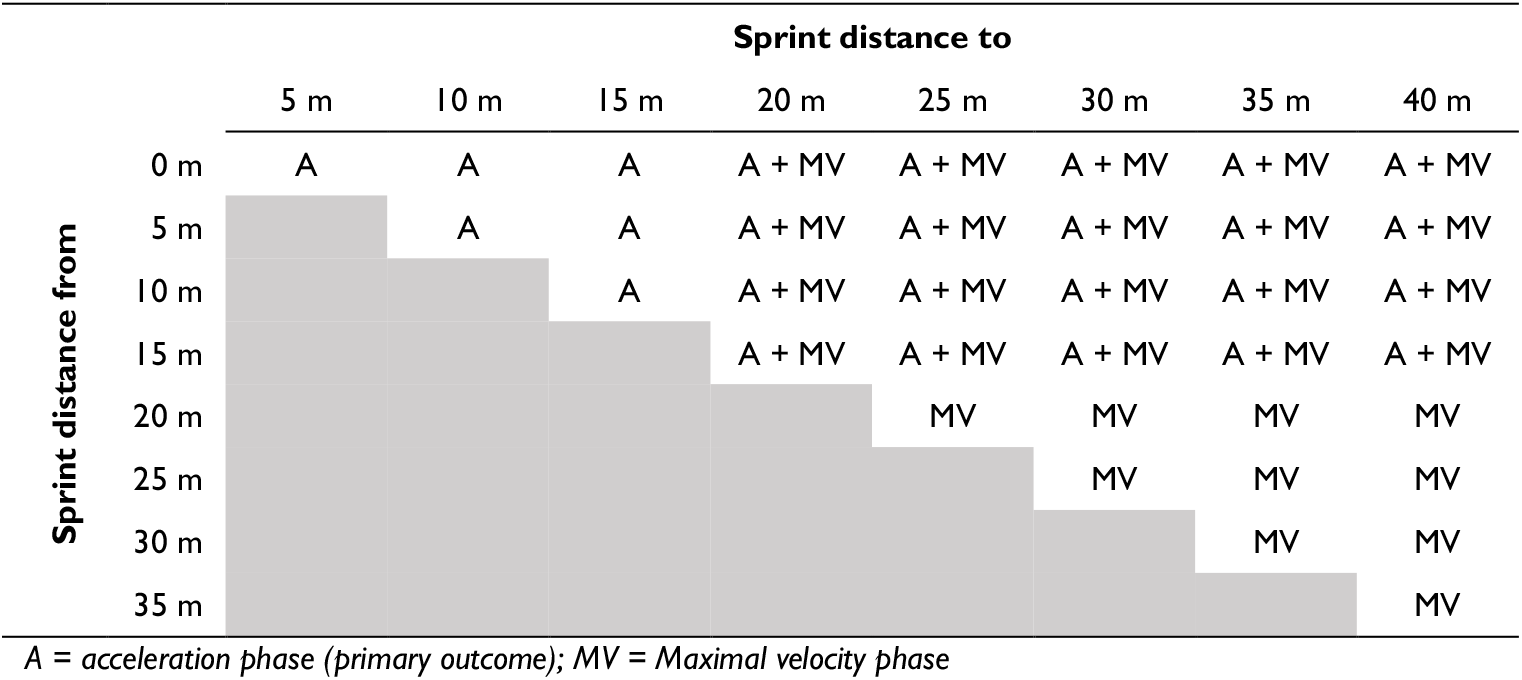
Split sprint distances and their outcome-designation

|  | Sprint distance to |  |  |  |  |  |  |  |
| --- | --- | --- | --- | --- | --- | --- | --- | --- |
|  | 5 m | 10 m | 15 m | 20 m | 25 m | 30 m | 35 m | 40 m |
| 0 m | A | A | A | A + MV | A + MV | A + MV | A + MV | A + MV |
| 5 m |  | A | A | A + MV | A + MV | A + MV | A + MV | A + MV |
| 10 m |  |  | A | A + MV | A + MV | A + MV | A + MV | A + MV |
| 15 m |  |  |  | A + MV | A + MV | A + MV | A + MV | A + MV |
| 20 m |  |  |  |  | MV | MV | MV | MV |
| 25 m |  |  |  |  |  | MV | MV | MV |
| 30 m |  |  |  |  |  |  | MV | MV |
| 35 m |  |  |  |  |  |  |  | MV |

A = acceleration phase (primary outcome); MV = Maximal velocity phase

**Table 2.**
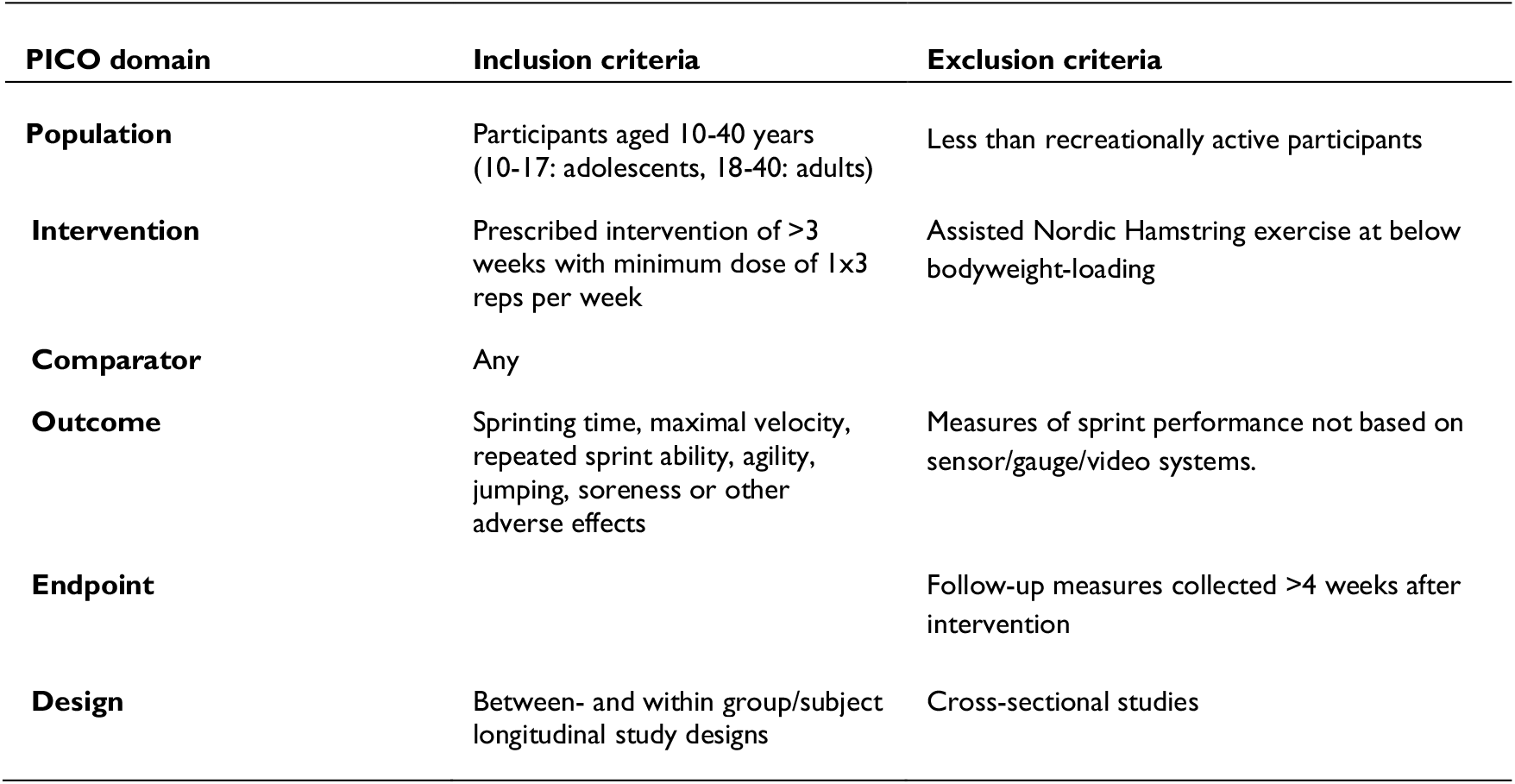
Short-form overview of key inclusion and exclusion criteria for eligible studies.

**Table 3.**
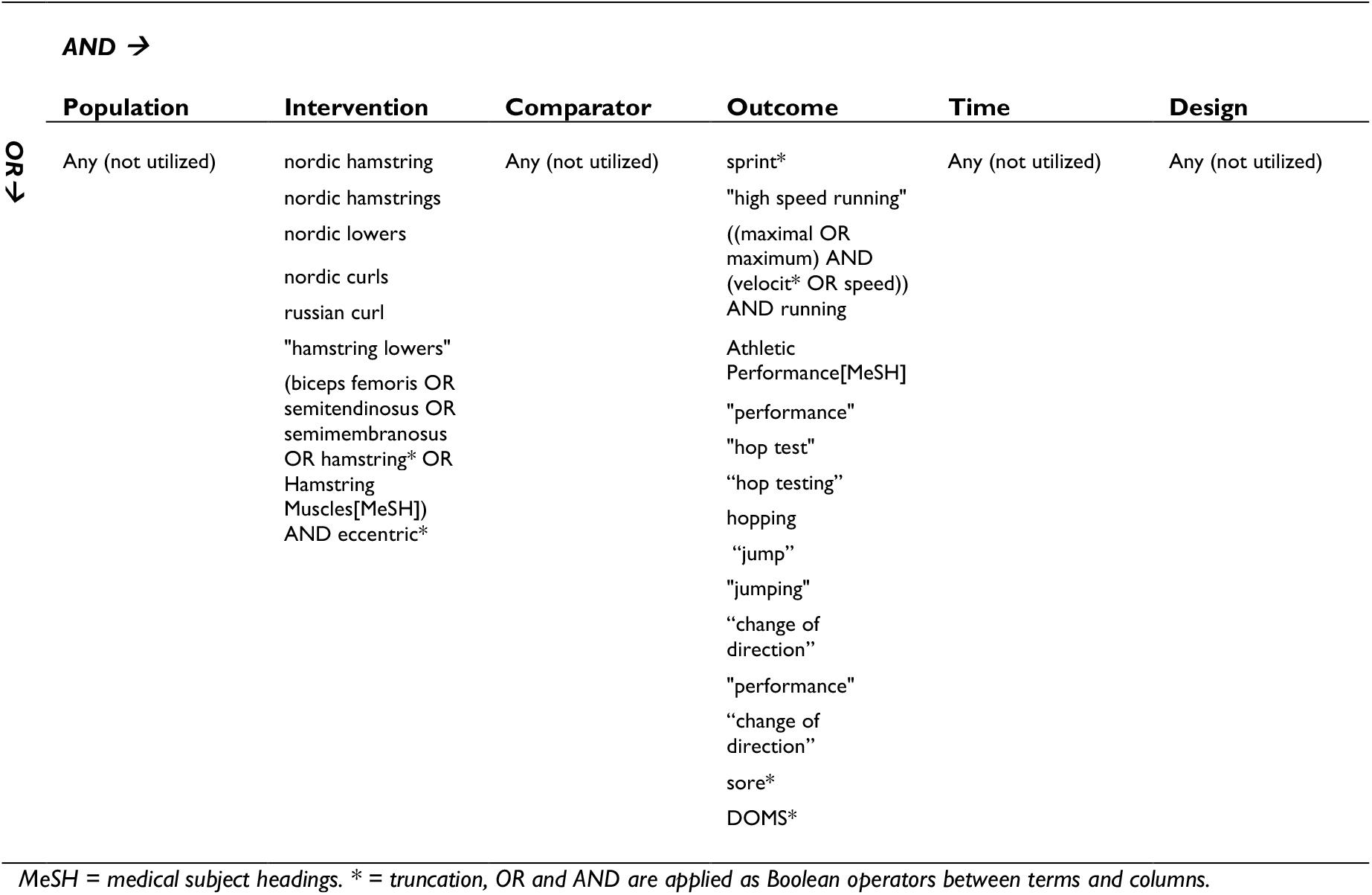
Search matrix and terms for PubMed.

Two reviewers (KK & LI) will independently agree on selection of eligible studies to include using Endnote (Clarivate Analytics) to remove duplicates and screen records. If a consensus cannot be achieved a third reviewer (BG) will be involved. A improved PRISMA-style flowchart^54^ will be presented, and we will provide a list of studies excluded on abstract or full-text level as a supplementary. Entries from some registries will also be immediately screened in full to assess possible eligibility as importation to reference manager is not possible (OSF, ISRCTN, SportRxiv, MedRxiv). The level of inter-rater agreement at the abstract-level between reviewers before consensus will be reported as percentage of agreement and unweighted kappa values. Along with the publication of this protocol as pre-print, we are contacting field experts through social media and email for suggestion or additional possible relevant studies for inclusion. Citations will be managed in Endnote.

### 2.4 Data extraction

Duplicate data extraction will be performed by three independent reviewers, with two reviewers per study (MF, LK, BG),^55^ using a piloted data extraction form based on Cochrane templates^56^ and Center for Evidence-Based Medicine guidelines.^57^ we have provided the un-populated form as a supplementary (Data Extraction Form.pdf). Data only presented as visualization will be extracted using a web-based tool (WebPlotDigitizer 4.2, 2019, https://automeris.io/WebPlotDigitizer). For extracting adverse events we will use a hybrid confirmatory-exploratory approach, in that we focus on soreness specifically and hereby pre-specified, and will also include any potential unrecognized or unspecified adverse events.^58^ For multiplicity of effect estimates, we will use an reductionist approach by including one estimate per analysis, per study-group, based on the follow criteria: 1) most recent (earliest follow-up measure) estimate, 2) intention-to-treat estimates, and 3) for sprint split times, the split closest to 20 meters.^59^ In cases of either missing data, or data extraction from visual reporting, we will contact corresponding authors by email and ask to either provide, or verify/correct data extraction values. For this, authors will be asked to supply data and study information through an electronic survey with the core data extraction items. After the initial email request, a reminder after 1 week, and contact by telephone (if telephone number is available and in service),^60^ none or the un-verified data extraction will be utilized. As a supplementary, we will list all studies included for which corresponding authors could not supply data or verify our visual extractions.

### 2.5 Critical appraisal

We will use Cochrane Risk of Bias Tool 2.0^41^ to evaluate risk of bias in randomized trials.^61^ The ROBINS-I tool will be used for non-randomized trials.^62^ Risk of Bias evaluation will be done once per study based on outcomes as prioritized in the outcome-section. For sprint-outcomes where several eligible sprint-distances are available, only the ones closest to the 20 m cut-off will be used as to not double-count participant-data. No study will be excluded based on the results of the critical appraisal, but this information will be used for the quality of cumulative evidence assessment using the Cochrane Collaborations GRADE criteria^63^ and for subgroup analysis of ‘low risk of bias’ studies.^30,64^ Rating the quality of individual studies rather than assessing risk of bias is inappropriate and will not be performed.^65,66^ Reviewers will perform duplicate independent bias assessment with the assistance of a machine learning system^67,68^ (MF, LK, & BG), and duplicate GRADE evaluation (KK & LI) and will achieve consensus in case of conflicting assessments by involving an additional reviewer (KK & BG). The level of inter-tester agreement between appraisers before consensus will be reported as percentage of agreement and unweighted kappa values. In sports- and exercise trials, blinding participants and personnel delivering exercise-based interventions are not feasible and we will therefore not consider domains pertaining to these blinding situations when assigning studies to “low risk of bias” subgroup analysis. Details from included studies pertaining to reporting of funding, conflict of interests, transparency, intention-to-treat analyses and pre-registration will be reported alongside general study characteristics.

### 2.6 Analyses

Our meta-analyses of individual study results will be performed with a random effects model using the Restricted Maximum Likelihood (REML) method to estimate a pooled mean of the overall effect with 95% confidence intervals, and the between study variance of effect sizes (Tau-Squared, T^2^). We will report estimated effect size and interpreted it as a standardized mean difference (SMD) calculated as Cohens’ d. However, we will use a Hedges’ g correction to account for small sized studies. Heterogeneity in the meta-analyses will be tested with Cochranes’ Q-test (a chi^2^-test).^66^ The percentage of variability in the effect estimates which is due to heterogeneity between the studies rather than chance will be calculated and interpreted using the I^2^ statistic.^66^ Causes for heterogeneity will be explored through subgroup analyses and meta-regression analyses of clinical and/or methodological covariates. Categorical covariates (e.g. age categories, playing-level, comparator, duration or volume of the Nordic Hamstring exercise) will be investigated with stratified meta-analysis and continuous covariates (e.g. intervention dose) will be investigated with meta-regression. A covariate is considered relevant and to explain heterogeneity in the overall analyses if inclusion of the covariate reduces the overall Tau-Squared. If a substantial clinical heterogeneity or high risk of bias is found, combining data in meta-analysis might be inappropriate, hence analysis will only be presented without total effect estimates, as firm conclusions on these will be inappropriate. Instead, we will present a brief narrative analysis, and the possible causes for heterogeneity will be debated in the discussion-section. No other qualitative syntheses are planned. Differences in estimated effects between relevant categorical subgroups will be analysed using meta-regression. For evaluating the risk of publication- and small study bias, a contour-enhanced funnel plot^69^ will be provided in addition to the Eggers test for Funnel-asymmetry and small study bias^70^ and Beggs test for publication bias.^71^ If relevant, reasons for funnel-asymmetry will be debated in the discussion-section. Results of the two primary outcomes will be presented in forest plots to allow for visual comparisons between studies. To allow for a more clinically relevant interpretation of the meta-analyses estimates, we will convert the pooled estimate from SMD to log (Odds Ratio), so we can estimate the odds ratio for becoming faster or slower and calculate the number needed to treat for 1 athlete to e.g. get faster. Furthermore, we will also convert the pooled estimate from SMD to an original scale (e.g. seconds), to enhance clinical interpretation of the results. All analyses will be performed in Stata 16 IC (StataCorp LLC, USA). All planned analyses are provided in table 4. Results will be emphasized based on effect size and confidence intervals, rather than statistical significance.

**Table 4.** Overview of planned meta-analyses.

| Primary analyses | Secondary analyses | Sensitivity and/or meta-regression analyses |
| --- | --- | --- |
| <20 meter sprint time (seconds) | ≥20 meter sprint time (seconds) | Only studies enrolling sub-elite or elite athletes |
|  | Maximum velocity (velocity and 20-40 m split times) | Only low risk of bias studies |
|  | Repeated sprint ability time | Only RCT studies |
|  | Agility test time pooled | Only adolescents (10-17 years) |
|  | All types of jumps pooled | Only adults (18-40 years) |
|  | Soreness | Only studies examining the Nordic Hamstring exercise without additive interventions |
|  | Acute effect on any sprint distance pooled | Only studies with other types of strength training as comparator |
|  |  | Only studies with specific sprint training as comparator |
|  |  | Only Nordic volume corresponding to FIFA 11+ dose or less weekly |
|  |  | Only Nordic volume corresponding to more than FIFA 11+ weekly |
|  |  | Only Nordic intervention of >4 weeks |
|  |  | Only Nordic intervention of ≤4 weeks |

*FIFA 11+ = a standardized warm-up program for football, including a progressive dose of the Nordic Hamstring exercise (2x5 reps, 3/week); RCT = Randomized controlled trial.*

## Data Availability

After publication, all data (populated data extraction forms, search files, supplementary analyses, statistical code, bias- and GRADE (Grading of Recommendations Assessment, Development and Evaluation) assessments, populated PRISMA checklist) will be shared as supplementary files with no restrictions, either on our institutional website, per journal repository or other open access repository (e.g. figshare.com).

## Funding and support

No specific funding were given for this study. No funding-or institutional body will have any control or involvement in data collection, analyses, interpretation, writing the report or decision to publish.

### Changelog from version 1.0 to 2.0 (04-JUNE-2021: before commencement of search)

- Added DOI
- Added soreness to search matrix
- Added specific trials registries
- Due to bugs and installation issues, SRA-helper and de-duplikator tools will not be utilized, and have been removed from the methods section
- Due to pandemical issues, the planned dates for carrying out the search has been postponed to June 8-16, 2020

### Changelog from version 2.0 to 3.0 (05-FEB-2021: screening started, next step is full-text assessments)

- Added PROSPERO registration ID.
- Added a sensitivity analysis: “Only studies with other types of strength training as comparator”, as this was mistakenly left out in previous drafts.
- Removed a sensitivity analysis from table 4: “Only studies with usual training or no intervention as comparator”, as this was in fact our primary analysis and was duplicated in table 4 mistakenly.
- Post hoc change: Added further details on the data extraction of grey literature from authors, as we have started this process earlier than anticipated. We are currently doing it before full-text assessments, rather than after data-extraction as anticipated.
- Post hoc change: Added details on screening of registry entries that was not possible to import into reference manager and was thus screen in full directly in the registry.

## Acknowledgement

The authors would like to thank Strength and Conditioning Coach Michael Myhre and Research Librarian Torben Jørgensen with help in choosing search terms and refining the search strategy.

## Author contributions

All authors contributed to the protocol. Contributions are visualized according to the CRediT framework (the Contributor Roles Taxonomy)^72^ in the following table.

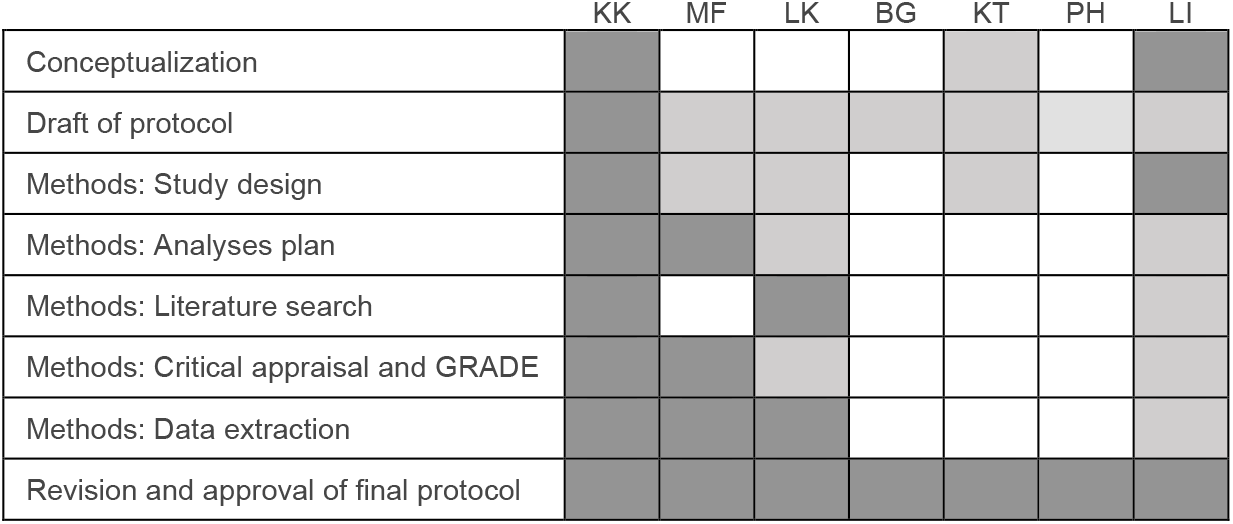

